# The burden of mortality due to injury in Cabo Verde, 2018

**DOI:** 10.1101/2022.11.21.22282603

**Authors:** Ngibo Mubeta Fernandes, Maria da Luz Lima Mendonça, Lara Ferrero Gomez

## Abstract

External causes continue to be one of the main causes of mortality in the world and Cabo Verde is no exception. Economic evaluations can be used to demonstrate the disease burden of public health problems such as injuries and external causes and support prioritization of interventions aimed at improving the health of the populations. The study objective was to estimate the indirect costs of premature mortality in 2018 due to injuries and other consequences of external causes in Cabo Verde. Years of potential life lost, years of potential productive life lost and human capital approach were used to estimate the burden and indirect costs of premature mortality in 2018. In 2018, 244 deaths were registered due to injury and other consequences of external causes. Males were responsible for 85.4% and 87.73% of years of potential life lost and years of potential productive life lost, respectively. The cost of productivity lost due to premature death caused by injury was 4,580,225.91 USD. The was social and economic burden due to trauma was significant. There is a need for more evidence on the burden of disease due to injuries and their consequences, to support the implementation of targeted multi-sectoral strategies and policies for the prevention, management, and reduction of costs due to injuries in Cabo Verde.

## Introduction

The world loses nearly five million people annually to trauma, poisoning and other external causes, which corresponds to 9% of global deaths [1]. Injuries continue to have significant weight on the global disease burden, and are among the main causes of morbidity and mortality in adolescents, young people and adults [2). In addition, injury represent a considerable public health problem, especially in developing countries, and are responsible for almost 90% of the world’s fatalities [3,4]. Furthermore, the burden of injuries and external causes of morbidity and mortality has a considerable social and economic impact. Road accidents were the main cause of death in young people and cause losses of up to 3% of the Gross Domestic Product (GDP) globally and up to 5% in low- and middle-income countries [5].

Despite being a considerable public health problem, injuries and their consequences are not prioritized on the world agenda [1,6]. Recognizing the magnitude of injuries and other consequences of external causes, goals have been set globally to reduce premature mortality rates from suicides and road accidents, strengthening prevention and treatment of important determinants of injuries and external causes by 2030 [5,7].

Injuries and the consequences of external causes are a major burden of disease globally, but mainly occur in developing countries, where few studies evaluating their economic impact and burden of diseases have been carried out [6]. Economic analyzes of interventions in the implementation of intervention policies to assess the results and costs of these interventions aimed at improving the health of the population are recommended [4,8] and used to demonstrate the disease burden in monetary terms [9–11].

Injuries continue to be one of the main causes of mortality and morbidity in the world, and Cabo Verde is no exception. In 2018, Cabo Verde recorded 2838 deaths, of which 244 were due to trauma, which represented a rate of five (5) deaths per 1000 inhabitants were the second leading cause of admissions in central hospitals in the country [12]. In addition, the country recognizes external causes as a public health problem that deserve the proper framework in the national health policy and a multisectoral approach to address this problem [13]. Cabo Verde is one of the Small Island Developing States (SIDS), and is located 450-500 km from the west coast of Africa and is characterized by scarce natural resources, recurrent drought, with a relatively small market economy. Cabo Verde is a lower middle-income country with a tourism-based economy and political stability and good governance have contributed to the economic success of the country [14]. The archipelago is made up of 10 islands and has a land surface of 4,033 square kilometers [15] and had an estimated population of 544,081 in 2018 [16].

The current study is the first carried out in Cabo Verde estimating the indirect costs of mortality due to injuries and consequences of external causes and demonstrating the socioeconomic burden.

## Materials and Methods

### General approach

The study is an observational, cross-sectional, a descriptive analysis of the epidemiological profile of premature mortality due to injury and a partial economic evaluation of the costs of premature mortality, adopting the socioeconomic perspective. The mortality data used, deaths from injuries were classified into two groups: external causes and injuries and consequences of external causes. For this study, and all deaths classified in both groups were considered.

### Source and data collection

The Integrated Surveillance and Response Service of the National Health Directorate of the Ministry of Health and Social Security (MSSS) made available mortality data for 2018. The data were anonymized, in order to guarantee the confidentiality of individual data. Deaths were filtered by age, sex, municipality and cause of death. All deaths classified with codes S00-T98 and V01-Y98 of the International Classification of Diseases and Related Health Problems, Tenth Revision (ICD-10) were extracted for analysis [17]. Cabo Verde mortality data are verified by the Ministry of Health, National Institute of Statistics, Notary and Identification to ensure data accuracy.

Descriptive data analysis was performed using Microsoft Excel 2016 and Statistical Package for Social Science (SPSS, version 22) and Mendeley Reference Management Software [18], was used to organize the bibliographic references.

### Estimation methods

Life expectancy and the human capital approach were used to estimate the burden and costs of premature mortality. The human capital approach was applied to estimate the value of potential lost productivity due to mortality due to injury [19]. To estimate the disease burden of premature mortality, measures of YPLL, YPPLL and CPL were used, [9,10,19,20]. These concepts estimate the average time a person would live if they did not die prematurely, taking into account the age limit (death/retirement) and the cause,[9]. YPLL estimates were based on the average value of life expectancy at birth in 2018 [21].

The YPLL were determined using the formula:

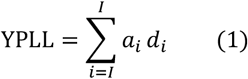

Where: *a*_*i*_= (73 −i−0.5) the number of years left to the stipulated age limit, *d*_*i*_ = the number of deaths in the age between i and i + 1.

YPLL rates (TYPLL) were stratified by municipality, sex, age group and cause of death.

Demographic projections were used to calculate the YPLL rates [16].

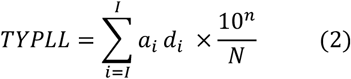

Where: N = number of people between 1 and 72 years of age in the population. Note: ai and di are already defined above.

To estimate the YPPLL, the age groups from 15 to 64 years old constituted the national workforce [22]

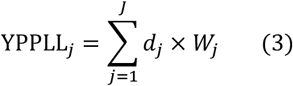

Where: *d*_*j*_ = average number of deaths for each age group W_*j*_=midpoint for retirement age in each age group =65-*j*-0.5 and *j*=are groups of 5 years for the entire productive population (15-64).

The YPPLL and the Gross Domestic Product (GDP) *per capita* of 3617.33 USD for the year 2018 [21), were used to estimate the CPL due to premature mortality using the following formula:

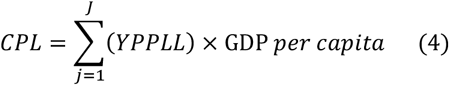

For this study it was assumed that the individual’s productivity did not change until retirement [9]. The present values of future costs were calculated by applying a discount rate of 4.5% practiced by the Bank of Cabo Verde [23]. In addition, a univariate sensitivity analysis was performed considering alternative discount rates (3% and 6%) to determine how different discount rate values will affect present productivity costs [8,24].

The present value was obtained using the formula:

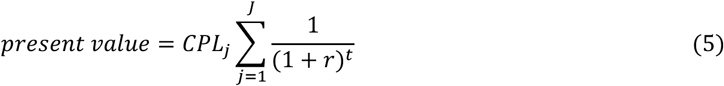

Where: *CPL* = future costs for each age group, *r* =discount rate, *t*=average point to reach retirement age for each age group and *j*=are groups of 5 years for the entire productive population (15-64).

### Ethical and legal considerations

This study was approved by the National Ethics Committee for Health Research (*CNEPS*) through resolution nº 33/2020 of May 28, 2020 and the National Commission for Data Protection (*CNPD*) through dispatch nº 183/2020. Being a retrospective study, no informed consent was required.

## Results

### Mortality profile

Cabo Verde recorded 244 deaths from injuries and external causes in 2018. Males were responsible for 84.4% of deaths. Regarding occupation, around 24.2% of deaths corresponded to elementary occupations Table 1.

**Table 1.** Distribution of deaths due to injury according to sociodemographic characteristics, Cape Verde, 2018.

| Variables | n (244) | % |
| --- | --- | --- |
| **Sex** |  |  |
| Male | 206 | 84.4 |
| Female | 38 | 15.6 |
| **marital status** |  |  |
| Single | 178 | 73.0 |
| Married/de facto union | 27 | 11.1 |
| Divorced / Separated | 6 | 2.5 |
| Widower | 8 | 3.3 |
| Unknown | 25 | 10.2 |
| **Profession** |  |  |
| Students | 8 | 3.9 |
| Specialists in intellectual and scientific activities | 1 | 0.5 |
| Intermediate level technicians and professionals | 7 | 3.4 |
| Personal protection services personnel | 7 | 3.4 |
| Farmers and skilled workers in agriculture and fisheries | 15 | 7.3 |
| Plant and machine operators and assembly workers | 7 | 3.4 |
| Elementary professions | 48 | 23.3 |
| Others | 57 | 27.7 |
| Unknown | 56 | 27.2 |

Regarding the specific causes of death, most deaths were due to traumatic head injuries (23.0%) and intentional self-harm (22.5%) in the observed population. The main causes of death among females were assaults (21.1%), followed by head injuries (18.4%) and intentionally self-inflicted injuries which accounted for 15.8% of deaths. Overall, the groups with the highest proportion of deaths were those between 20 to 24, 25 to 29 and 35 to 39, which represent 34.4% of the cases registered in 2018 Table 2.

**Table 2.** Distribution of deaths by sex and cause, Cabo Verde, 2018.

| Causes of death | Male | % | Female | % | Total | % |
| --- | --- | --- | --- | --- | --- | --- |
| Injuries to the head | 49 | 23.8 | 7 | 18.4 | 56 | 23.0 |
| Injuries to the neck | 4 | 1.9 | 0 | 0.0 | 4 | 1.6 |
| Injuries to the abdomen, lower back, lumbar spine and pelvis | 1 | 0.5 | 0 | 0.0 | 1 | 0.4 |
| Injuries to the hip and thigh | 2 | 1.0 | 4 | 10.5 | 6 | 2.5 |
| Injuries involving multiple body regions | 33 | 16.0 | 5 | 13.2 | 38 | 15.6 |
| Injuries to unspecified part of trunk, limb, or body region | 1 | 0.5 | 0 | 0.0 | 1 | 0.4 |
| Burns and corrosions | 2 | 1.0 | 3 | 7.9 | 5 | 2.0 |
| Poisoning by drugs, medicaments and biological substances | 1 | 0.5 | 0 | 0.0 | 1 | 0.4 |
| Toxic effects of substances of chiefly non-medicinal origin | 3 | 1.5 | 2 | 5.3 | 5 | 2.0 |
| Other and unspecified effects of external causes | 4 | 1.9 | 1 | 2.6 | 5 | 2.0 |
| Certain early complications of trauma | 6 | 2.9 | 0 | 0.0 | 6 | 2.5 |
| Accidental drowning and submersion | 23 | 11.2 | 1 | 2.6 | 24 | 9.8 |
| Other accidental threats to breathing | 5 | 2.4 | 1 | 2.6 | 6 | 2.5 |
| Intentional self-harm | 49 | 23.8 | 6 | 15.8 | 55 | 22.5 |
| Assault | 23 | 11.2 | 8 | 21.1 | 31 | 12.7 |
| **Total** | **206** | **100** | **38** | **100** | **244** | **100** |

### Years of potential life lost

Of the 244 deaths reported due to trauma, 211 (86.5%) were in individuals aged between 1 and 72 years old and this accounted 7222 YPLL and on average, 34.2 YPLL were lost per death. Males lost on average of 33.7 YPLL and females lost 37.6 YPLL per death. The age range 20 to 39 years old, accounted for 60.3% or 4357 YPLL, the same trend was observed in the male population, where 63.3% or 3906 YPLL. For females, the age group 1 to 4 years contributed 20% (210 YPLL) of the total YPLL in this group, compared to 1.9% (140) for males. Approximately 14 years were lost per 1000 inhabitants between 1 and 72 in 2018. Males and females had rates of 23.6 and 4.2 potential years of life lost per 1000 inhabitants, respectively Table 3.

**Table 3.** Years of potential life lost, proportions, means and rates per 1000 inhabitants by sex and age group, Cabo Verde, 2018.

| Age group | Male |  |  | Female |  |  | Total |  |  |
| --- | --- | --- | --- | --- | --- | --- | --- | --- | --- |
|  | YPLL | % | TYPLL | YPLL | % | TYPLL | YPLL | % | TYPLL |
| 1 to 4 | 140 | 1.9 | 6.5 | 210 | 2.9 | 10.3 | 350 | 4.8 | 8.4 |
| 5 to 9 | 262 | 3.6 | 10.4 | 66 | 0.9 | 2.7 | 328 | 4.5 | 6.6 |
| 10 to 14 | 242 | 3.4 | 9.4 | 0.0 | 0.0 | 0.0 | 242 | 3.4 | 4.7 |
| 15 to 19 | 111 | 1.5 | 4.6 | 111 | 1.5 | 4.7 | 222 | 3.1 | 4.6 |
| 20 to 24 | 1212 | 16.8 | 47.5 | 152 | 2.1 | 6.2 | 1364 | 18.9 | 27.3 |
| 25 to 29 | 865 | 12.0 | 30.1 | 228 | 3.2 | 8.7 | 1092 | 15.1 | 19.9 |
| 30 to 34 | 729 | 10.1 | 27.1 | 0.0 | 0.0 | 0.0 | 729 | 10.1 | 14.5 |
| 35 to 39 | 1101 | 15.2 | 50.0 | 71 | 1.0 | 3.9 | 1172 | 16.2 | 29.0 |
| 40 to 44 | 610 | 8.4 | 36.7 | 31 | 0.4 | 2.1 | 641 | 8.9 | 20.5 |
| 45 to 49 | 230 | 3.2 | 17.1 | 77 | 1.1 | 6.0 | 306 | 4.2 | 11.7 |
| 50 to 54 | 246 | 3.4 | 20.4 | 21 | 0.3 | 1.6 | 267 | 3.7 | 10.7 |
| 55 to 59 | 279 | 3.9 | 29.5 | 47 | 0.6 | 4.3 | 326 | 4.5 | 16.1 |
| 60 to 64 | 105 | 1.5 | 18.4 | 42 | 0.6 | 5.1 | 147 | 2.0 | 10.5 |
| 65 to 69 | 33 | 0.5 | 10.2 | 0 | 0.0 | 0.0 | 33 | 0.5 | 4.0 |
| 70 to 72 | 6 | 0.1 | 5.2 | 0 | 0.0 | 0.0 | 6 | 0.1 | 2.1 |
| **Total** | **6170** | **85.4** | **23.6** | **1053** | **14.6** | **4.2** | **7222** | **100** | **14.0** |

### The geographic distribution of YPLL

The distribution of YPLL due to injury varied between regions. The municipality of Praia contributed to the highest proportion of YPLL with 27.4%, followed by the island of São Vicente with (13.6%) and Santa Catarina with (10.8%). The municipality of Brava had the highest rate of 34.5 YPLL per 1000 inhabitants and the lowest rate was recorded in the municipality of São Lourenço dos Órgãos with 5.5 YPLL per 1000 inhabitants. The municipalities of São Domingos and Ribeira Grande Santiago revealed the highest averages of losses of 46.6 and 45.5 years per death and victims died on average, at 26.4 and 27.5 years, respectively. The municipality of Santa Catarina de Fogo had the lowest mean per death of 18.8 and, subsequently, the highest mean per death, of 54.2 years. The mean age at death from external causes in 2018 was 38.8 years Table 4.

**Table 4.** Years of potential life lost, proportions, means and rates per 1000 inhabitants by municipalities, Cabo Verde, 2018.

| Municipality | YPLL | % | TYPLL | Mean | Mean age at death |
| --- | --- | --- | --- | --- | --- |
| Brava | 178 | 2.5 | 34.5 | 29.7 | 43.3 |
| Paul | 147 | 2.0 | 28.3 | 36.8 | 36.3 |
| Tarrafal de São Nicolau | 122 | 1.7 | 25.3 | 30.5 | 42.5 |
| Ribeira Brava | 152 | 2.1 | 24.0 | 38.0 | 35.0 |
| Porto Novo | 377 | 5.2 | 23.8 | 29.0 | 44.0 |
| Tarrafal | 346 | 4.8 | 20.3 | 34.6 | 38.4 |
| Santa Catarina | 782 | 10.8 | 17.9 | 34.0 | 39.0 |
| São Salvador do Mundo | 132 | 1.8 | 16.5 | 43.8 | 29.2 |
| Boavista | 248 | 3.4 | 14.5 | 41.3 | 31.8 |
| Ribeira Grande | 204 | 2.8 | 14.0 | 29.1 | 43.9 |
| São Domingos | 187 | 2.6 | 14.0 | 46.6 | 26.4 |
| São Miguel | 177 | 2.5 | 13.4 | 44.3 | 28.8 |
| Praia | 1978 | 27.4 | 12.7 | 42.1 | 30.9 |
| São Vincent | 982 | 13.6 | 12.5 | 30.7 | 42.3 |
| Ribeira Grande de Santiago | 91 | 1.3 | 11.4 | 45.5 | 27.5 |
| Santa Catarina do Fogo | 57 | 0.8 | 11.4 | 18.8 | 54.2 |
| Maio | 76 | 1.1 | 11.2 | 38.0 | 35.0 |
| São Felipe | 219 | 3 | 11.2 | 27.4 | 45.6 |
| Sal | 368 | 5.1 | 10.0 | 28.3 | 44.7 |
| Santa Cruz | 188 | 2.6 | 7.6 | 31.3 | 41.7 |
| Mosteiros | 66 | 0.9 | 7.5 | 33.0 | 40.0 |
| São Lourenço dos Órgãos | 36 | 0.5 | 5.5 | 35.5 | 37.5 |
| Others | 114 | 1.6 | - | 16.2 | 56.8 |
| **Cabo Verde** | **7222** | **100** | **14.0** | **34.2** | **38.8** |

### Years of potential life lost by cause of death

Causes due to intentional self-harm contributed the largest proportion of the total YPLL (25.8% ;1862) and similarly, for males and was responsible for 26.6%;(1639). The largest contributor of YPLL for females were deaths from assault and accounted for 29.4% (309) of total YPLL among females. Regarding the mean YPLL per death, deaths from toxic effects of substances of essentially non-medicinal origin had the highest mean of 53.5 YPLL for both sexes and the same group of causes had the highest mean among males, 45 YPLL/death while for females, the highest average was 70 YPLL per deaths in the groups “causes from other effects and unspecified effects of external causes and toxic effects of substances of essentially non-medicinal origin” Table 5.

**Table 5.**
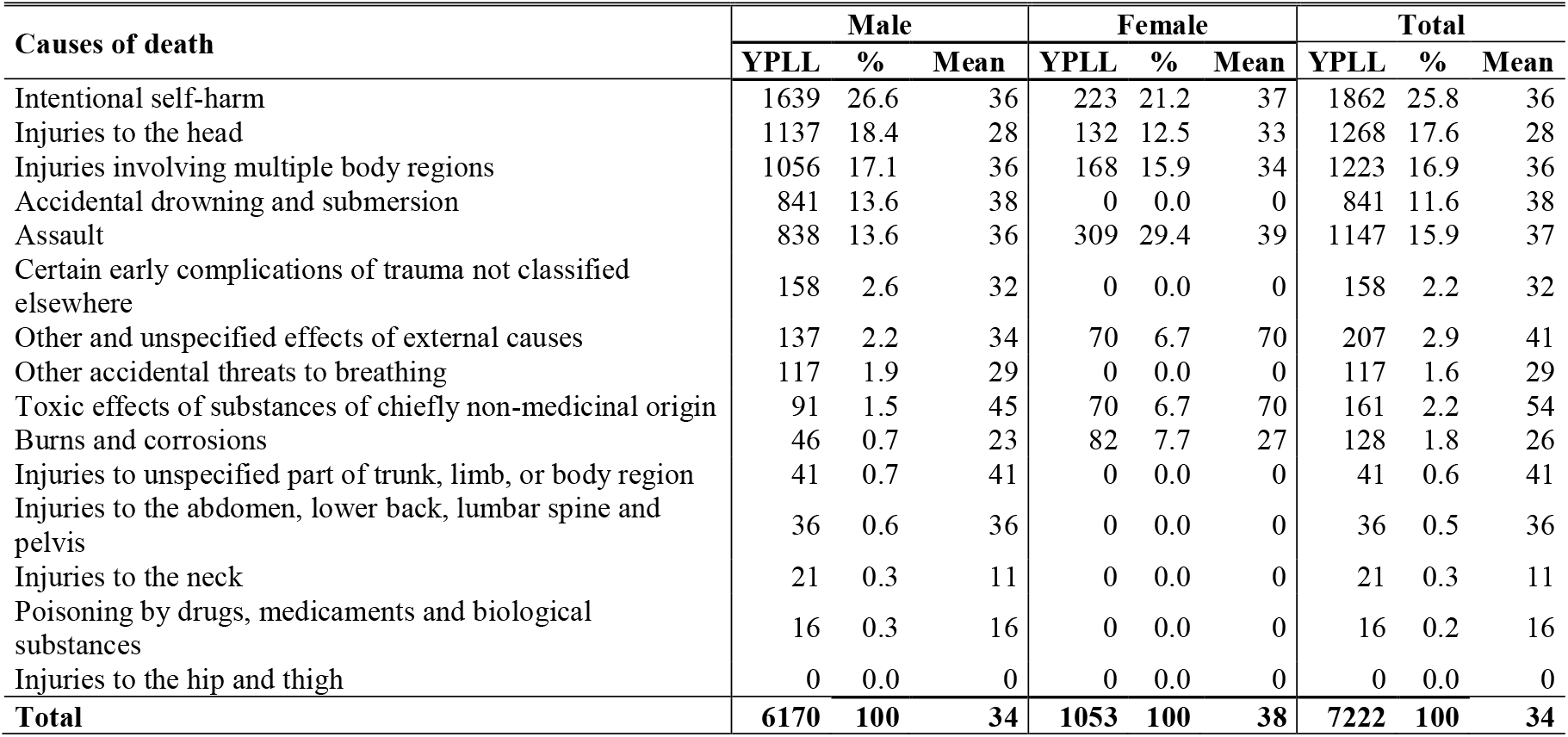
Years of potential life lost, proportions and means by causes, Cabo Verde, 2018.

### Years of potential productive life lost

Overall, 187 deaths occurred in individuals aged between 15 to 64 years and resulted in a total of 4768 YPPLL. Males and females, accounted for 87.7% (4183) and 12.3% (585) of total YPPLL, respectively. The highest proportions of YPPLL were observed in the age group 20 to 24 years and similarly for males and corresponded to 24.1% (1148), 24.4% (1020), respectively, while for females, the highest percentage was observed in the 25 to 29 age group (32.1%; 188). For both sexes, the YPPLL rate was 13.2 per 1000 inhabitants, 23 per 1000 inhabitants for males and 3 per 1000 inhabitants of the female population Table 6.

**Table 6.** Years of potential productive life lost, proportions, means and rates per 1000 inhabitants by sex and age group, Cabo Verde, 2018.

| Age group | Male |  |  | Female |  |  | Total |  |  |
| --- | --- | --- | --- | --- | --- | --- | --- | --- | --- |
|  | YPPLL | % | TYPPLL | YPPLL | % | TYPPLL | YPPLL | % | TYPPLL |
| 15 to 19 | 95 | 2.3 | 3.9 | 95 | 16.2 | 4.0 | 190 | 4 | 4.0 |
| 20 to 24 | 1020 | 24.4 | 40.0 | 128 | 21.9 | 5.2 | 1148 | 24.1 | 22.9 |
| 25 to 29 | 713 | 17.0 | 24.8 | 188 | 32.1 | 7.2 | 900 | 18.9 | 16.4 |
| 30 to 34 | 585 | 14.0 | 21.8 | 0 | 0.0 | 0.0 | 585 | 12.3 | 11.6 |
| 35 to 39 | 853 | 20.4 | 38.7 | 55 | 9.4 | 3.0 | 908 | 19 | 22.5 |
| 40 to 44 | 450 | 10.8 | 27.1 | 23 | 3.9 | 1.5 | 473 | 9.9 | 15.1 |
| 45 to 49 | 158 | 3.8 | 11.8 | 53 | 9.1 | 4.1 | 210 | 4.4 | 8.0 |
| 50 to 54 | 150 | 3.6 | 12.4 | 13 | 2.2 | 1.0 | 163 | 3.4 | 6.5 |
| 55 to 59 | 135 | 3.2 | 14.3 | 23 | 3.9 | 2.1 | 158 | 3.3 | 7.8 |
| 60 to 64 | 25 | 0.6 | 4.4 | 10 | 1.7 | 1.2 | 35 | 0.7 | 2.5 |
| **Total** | **4183** | **100** | **23.0** | **585** | **100** | **3.0** | **4768** | **100** | **13.2** |

### The geographic distribution of YPPLL

Of the 4768 YPPLL of injuries and external causes calculated, the municipality of Praia had the highest proportion, around 28.9% (1380), followed by the island of São Vicente with 12.9% (615) and the municipality of Santa Catarina with 10.2% (485). The geographic distribution of YPPLL rates was higher in the municipality of Brava with 37.9 per 1000 inhabitants, followed by the municipalities of the island of São Nicolau with 27.5 (Tarrafal) and 26.8 (Ribeira Brava). The municipalities of Ribeira Grande de Santiago and São Miguel recorded the highest losses, on average, of 37.5 and 36.3 YPPLL per death, respectively Table 7.

**Table 7.** Years of potential productive life lost, proportions, means and rates per 1000 inhabitants by municipalities, Cabo Verde, 2018.

| Municipality | YPPLL | % | Rate | Mean | Mean age at death |
| --- | --- | --- | --- | --- | --- |
| Boavista | 138 | 2.9 | 10.9 | 27.5 | 38 |
| Brava | 130 | 2.7 | 37.9 | 21.7 | 43 |
| Maia | 8 | 0.2 | 1.6 | 7.5 | 58 |
| Mosteiros | 50 | 1.0 | 8.7 | 25.0 | 40 |
| Others | 58 | 1.2 | 0.2 | 8.2 | 57 |
| Paul | 63 | 1.3 | 17.6 | 20.8 | 44 |
| Porto Novo | 278 | 5.8 | 25.3 | 25.2 | 40 |
| Praia | 1380 | 28.9 | 12.5 | 32.9 | 32 |
| Ribeira Brava | 120 | 2.5 | 26.8 | 30.0 | 35 |
| Ribeira Grande | 148 | 3.1 | 14.7 | 21.1 | 44 |
| Ribeira Grande de Santiago | 75 | 1.6 | 13.8 | 37.5 | 28 |
| Sal | 273 | 5.7 | 10.5 | 24.8 | 40 |
| Santa Catarina | 485 | 10.2 | 15.9 | 24.3 | 41 |
| Santa Catarina do Fogo | 33 | 0.7 | 10.3 | 10.8 | 54 |
| Santa Cruz | 140 | 2.9 | 8.6 | 23.3 | 42 |
| São Domingos | 93 | 1.9 | 10.4 | 30.8 | 34 |
| São Filipe | 158 | 3.3 | 12.3 | 22.5 | 43 |
| São Lourenço dos Órgãos | 28 | 0.6 | 6.2 | 27.5 | 38 |
| São Miguel | 145 | 3.0 | 16.4 | 36.3 | 29 |
| São Salvador do Mundo | 50 | 1.0 | 9.4 | 25.0 | 40 |
| Saint Vincent | 615 | 12.9 | 10.7 | 22.0 | 43 |
| Tarrafal | 215 | 4.5 | 18.8 | 26.9 | 38 |
| Tarrafal de São Nicolau | 90 | 1.9 | 27.5 | 22.5 | 43 |
| **Cabo Verde** | **4768** | **100** | **13.2** | **25.5** | **40** |

### Years of potential productive life lost by cause of death

With regards to causes, for both sexes, intentional self-harm contributed 30.6% of YPPLL, followed by assaults with 19%, injuries involving multiple body regions with 17% and head injuries with 14%. On average, fatal victim of assaults recorded the highest mean YPPLL of 31, 32 and 31 for both sexes, for males and females, respectively. The overall mean YPPLL per death was 25, 26 YPPLL per death for males and 24 YPPLL for females. On average, males died at 39.3 years and females at 40.6 years Table 8.

**Table 8.** Years of potential productive life lost, proportions and means by causes, Cabo Verde, 2018.

| Causes of death | Male |  |  | Female |  |  | Total |  |  |
| --- | --- | --- | --- | --- | --- | --- | --- | --- | --- |
|  | YPPLL | % | Mean | YPPLL | % | Mean | YPPLL | % | Mean |
| Intentional self-harm | 1283 | 30.7 | 30 | 175 | 29.9 | 29 | 1458 | 30.6 | 30 |
| Assault | 663 | 15.8 | 32 | 245 | 41.9 | 31 | 908 | 19.0 | 31 |
| Injuries involving multiple body regions | 725 | 17.3 | 28 | 70 | 12 | 18 | 795 | 16.7 | 27 |
| Injuries to the head | 655 | 15.7 | 19 | 38 | 6.4 | 13 | 693 | 14.5 | 19 |
| Accidental drowning and submersion | 435 | 10.4 | 24 | 0 | 0.0 | 0 | 435 | 9.1 | 24 |
| Certain early complications of trauma not classified elsewhere | 118 | 2.8 | 24 | 0 | 0.0 | 0 | 118 | 2.5 | 24 |
| Other and unspecified effects of external causes | 105 | 2.5 | 26 | 0 | 0.0 | 0 | 105 | 2.2 | 26 |
| Burns and Corrosions | 30 | 0.7 | 15 | 58 | 9.8 | 19 | 88 | 1.8 | 18 |
| Other accidental threats to breathing | 85 | 2.0 | 21 | 0 | 0.0 | 0 | 85 | 1.8 | 21 |
| injuries to unspecified part of trunk, limb, or body region | 33 | 0.8 | 33 | 0 | 0.0 | 0 | 33 | 0.7 | 33 |
| Injuries to the abdomen, lower back, lumbar spine and pelvis | 28 | 0.7 | 28 | 0 | 0.0 | 0 | 28 | 0.6 | 28 |
| Toxic effects of substances of chiefly non-medicinal origin | 13 | 0.3 | 13 | 0 | 0.0 | 0 | 13 | 0.3 | 13 |
| Poisoning by drugs, medicaments and biological substances | 8 | 0.2 | 8 | 0 | 0.0 | 0 | 8 | 0.2 | 8 |
| Injuries to the neck | 5 | 0.1 | 3 | 0 | 0.0 | 0 | 5 | 0.1 | 3 |
| Injuries to the hip and thigh | 0 | 0 | 0 | 0 | 0.0 | 0 | 0 | 0.0 | 0 |
| **Total** | **4183** | **100** | **26** | **585** | **100** | **24** | **4768** | **100** | **25** |

### Estimated costs of productivity lost

The estimated cost of productivity lost due to premature death attributable to injuries and external causes was 4,580,225.91 USD. Males contributed 87.7% (4,053,833.79 USD) of the total CPL in 2018. The average CPL per death 24 493.19 USD for both sexes, 24,870.15 USD for males and 21 933.01 USD for females. Approximately 21% (957,146.98USD) of the total CPL was attributed to individuals between 35 to 39 years for both sexes, and 22%; (899,138.07 USD), for males, while for females, the highest proportion of CPL (24.2%; 127,341.62 USD) was observed in the 25-29 age group Tables 9.

**Table 9.**
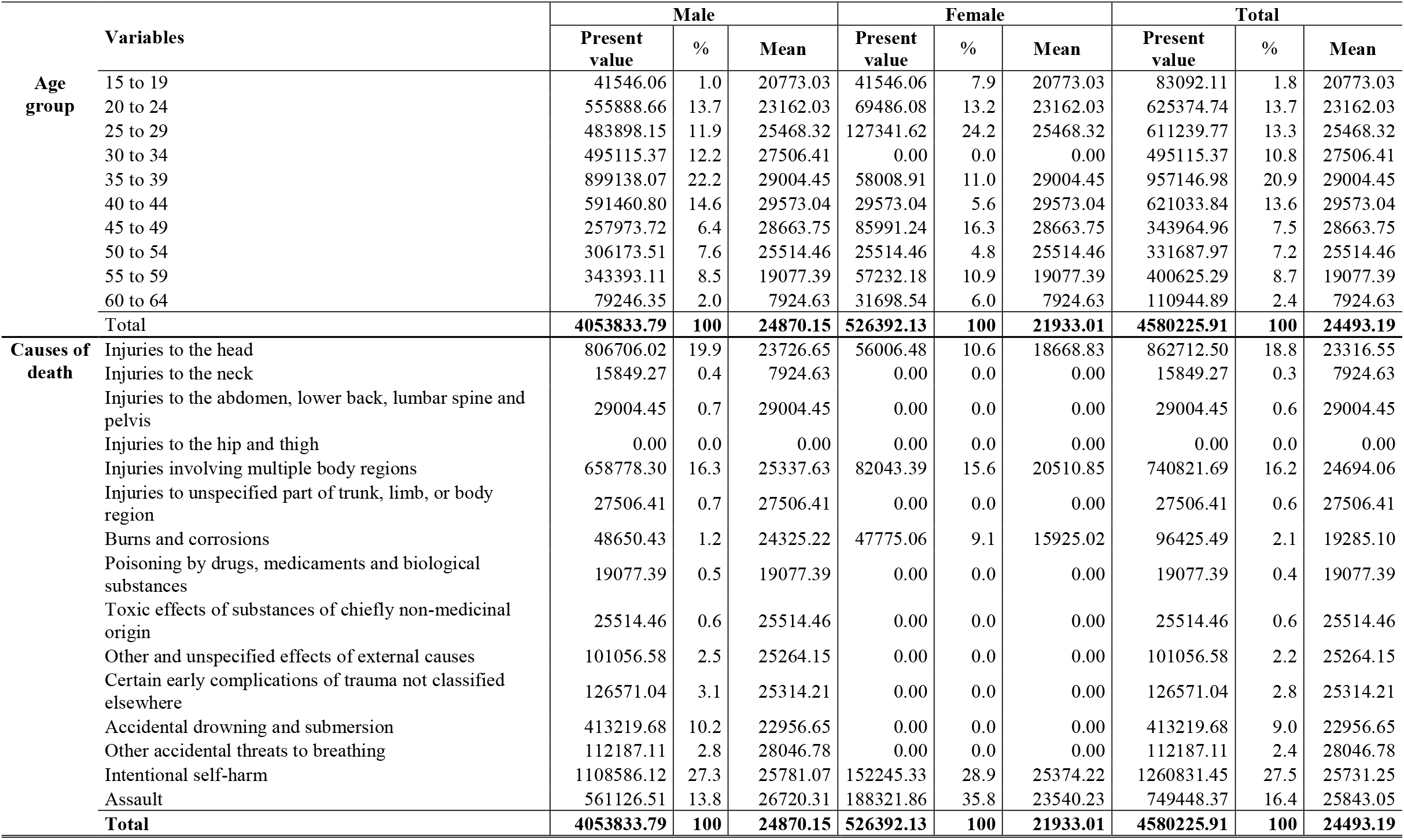
Estimated Costs of Lost Productivity by sex, age group and cause, Cabo Verde, 2018.

The principal drivers of CPL by specific causes were intentional self-harm, which was responsible for 27.5% (1,260,831.45 USD) of the total losses, followed by head injuries that contributed for 18.8% (862,712.50 USD). Furthermore, injuries to the abdomen, back, lumbar spine and pelvis had the highest mean loss per death, amounting to 29,004.45 USD. The same cause had the highest mean among males of 29,004.45 USD, while for females, the highest mean per death was attributed to the group intentionally self-inflicted injury in the amount of 25,374.22 USD followed by the cause “aggression” in the amount of 23540.23 USD per death Table 9.

A univariate sensitivity analysis was conducted to assess the effects of varying the discount rates. The analysis, at 3% was worth the total CPL 6,900,400.84 USD and 3,140,890.80 USD at 6%, with an average CPP per death of 36,900.54 USD and 16,796.21 USD, respectively. At 3%, the highest proportion of the loss which corresponded to 20.8% (1,434,803.65 USD) of the total CPL, was observed in the 35 to 39 age group and the highest proportion of loss in the amount of 1,976,502.46 USD was attributed to the “intentional self-harm” group. At 6%, proportions of CPL by age group and cause did not differ significantly, and the total CPL value was approximately 50% lower than the total CPP value at 3% Tables 10 and 11.

**Table 10.**
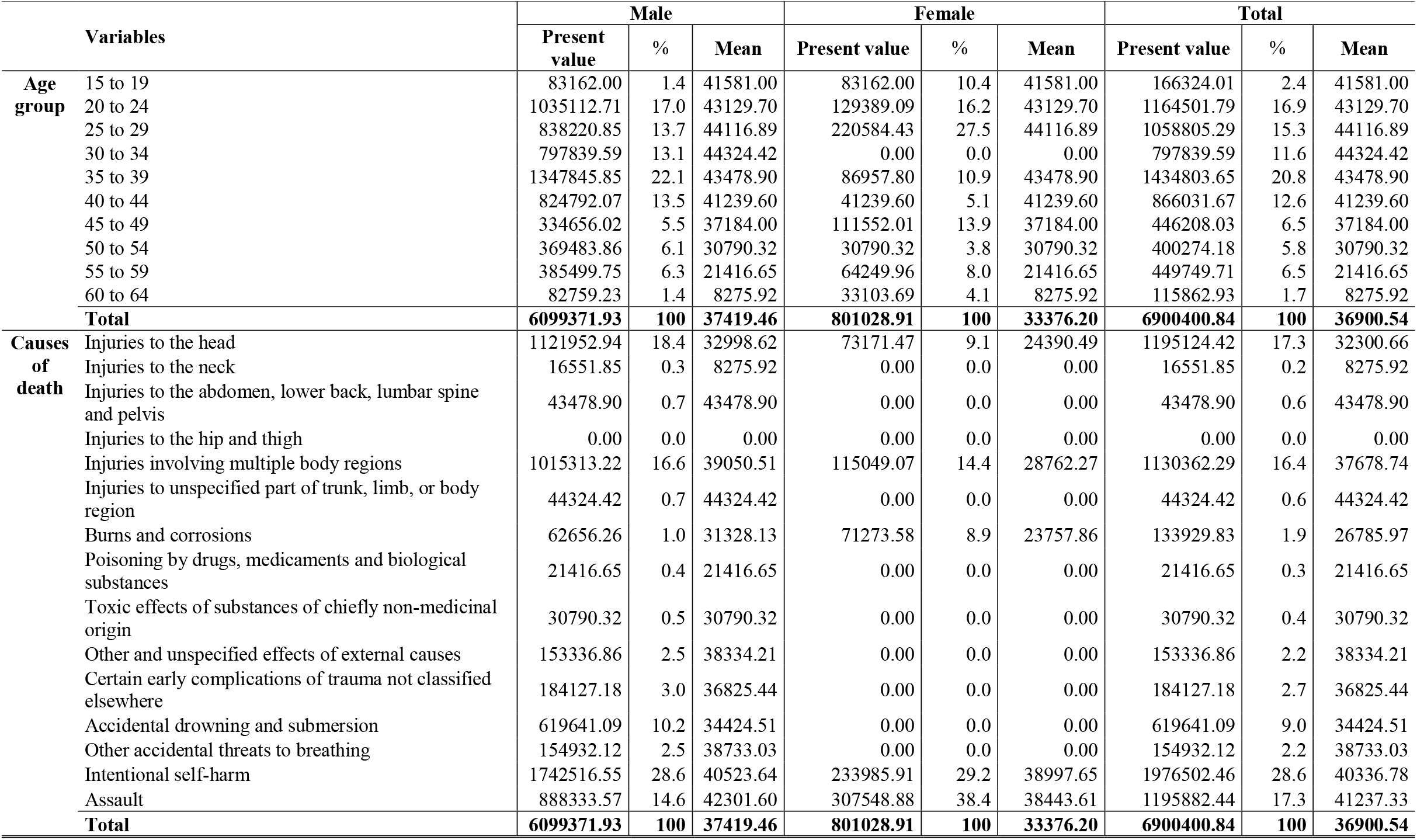
Sensitivity Analysis of Estimated Lost Productivity Costs by sex, age group and cause at 3%, Cabo Verde, 2018.

| Variables |  | Male |  |  | Female |  |  | Total |  |  |
| --- | --- | --- | --- | --- | --- | --- | --- | --- | --- | --- |
|  |  | Present value | % | Mean | Present value | % | Mean | Present value | % | Mean |
| **Age group** | 15 to 19 | 83162.00 | 1.4 | 41581.00 | 83162.00 | 10.4 | 41581.00 | 166324.01 | 2.4 | 41581.00 |
|  | 20 to 24 | 1035112.71 | 17.0 | 43129.70 | 129389.09 | 16.2 | 43129.70 | 1164501.79 | 16.9 | 43129.70 |
|  | 25 to 29 | 838220.85 | 13.7 | 44116.89 | 220584.43 | 27.5 | 44116.89 | 1058805.29 | 15.3 | 44116.89 |
|  | 30 to 34 | 797839.59 | 13.1 | 44324.42 | 0.00 | 0.0 | 0.00 | 797839.59 | 11.6 | 44324.42 |
|  | 35 to 39 | 1347845.85 | 22.1 | 43478.90 | 86957.80 | 10.9 | 43478.90 | 1434803.65 | 20.8 | 43478.90 |
|  | 40 to 44 | 824792.07 | 13.5 | 41239.60 | 41239.60 | 5.1 | 41239.60 | 866031.67 | 12.6 | 41239.60 |
|  | 45 to 49 | 334656.02 | 5.5 | 37184.00 | 111552.01 | 13.9 | 37184.00 | 446208.03 | 6.5 | 37184.00 |
|  | 50 to 54 | 369483.86 | 6.1 | 30790.32 | 30790.32 | 3.8 | 30790.32 | 400274.18 | 5.8 | 30790.32 |
|  | 55 to 59 | 385499.75 | 6.3 | 21416.65 | 64249.96 | 8.0 | 21416.65 | 449749.71 | 6.5 | 21416.65 |
|  | 60 to 64 | 82759.23 | 1.4 | 8275.92 | 33103.69 | 4.1 | 8275.92 | 115862.93 | 1.7 | 8275.92 |
| **Total** |  | **6099371.93** | **100** | **37419.46** | **801028.91** | **100** | **33376.20** | **6900400.84** | **100** | **36900.54** |
| **Causes of death** | Injuries to the head | 1121952.94 | 18.4 | 32998.62 | 73171.47 | 9.1 | 24390.49 | 1195124.42 | 17.3 | 32300.66 |
|  | Injuries to the neck | 16551.85 | 0.3 | 8275.92 | 0.00 | 0.0 | 0.00 | 16551.85 | 0.2 | 8275.92 |
|  | Injuries to the abdomen, lower back, lumbar spine and pelvis | 43478.90 | 0.7 | 43478.90 | 0.00 | 0.0 | 0.00 | 43478.90 | 0.6 | 43478.90 |
|  | Injuries to the hip and thigh | 0.00 | 0.0 | 0.00 | 0.00 | 0.0 | 0.00 | 0.00 | 0.0 | 0.00 |
|  | Injuries involving multiple body regions | 1015313.22 | 16.6 | 39050.51 | 115049.07 | 14.4 | 28762.27 | 1130362.29 | 16.4 | 37678.74 |
|  | Injuries to unspecified part of trunk, limb, or body region | 44324.42 | 0.7 | 44324.42 | 0.00 | 0.0 | 0.00 | 44324.42 | 0.6 | 44324.42 |
|  | Burns and corrosions | 62656.26 | 1.0 | 31328.13 | 71273.58 | 8.9 | 23757.86 | 133929.83 | 1.9 | 26785.97 |
|  | Poisoning by drugs, medicaments and biological substances | 21416.65 | 0.4 | 21416.65 | 0.00 | 0.0 | 0.00 | 21416.65 | 0.3 | 21416.65 |
|  | Toxic effects of substances of chiefly non-medicinal origin | 30790.32 | 0.5 | 30790.32 | 0.00 | 0.0 | 0.00 | 30790.32 | 0.4 | 30790.32 |
|  | Other and unspecified effects of external causes | 153336.86 | 2.5 | 38334.21 | 0.00 | 0.0 | 0.00 | 153336.86 | 2.2 | 38334.21 |
|  | Certain early complications of trauma not classified elsewhere | 184127.18 | 3.0 | 36825.44 | 0.00 | 0.0 | 0.00 | 184127.18 | 2.7 | 36825.44 |
|  | Accidental drowning and submersion | 619641.09 | 10.2 | 34424.51 | 0.00 | 0.0 | 0.00 | 619641.09 | 9.0 | 34424.51 |
|  | Other accidental threats to breathing | 154932.12 | 2.5 | 38733.03 | 0.00 | 0.0 | 0.00 | 154932.12 | 2.2 | 38733.03 |
|  | Intentional self-harm | 1742516.55 | 28.6 | 40523.64 | 233985.91 | 29.2 | 38997.65 | 1976502.46 | 28.6 | 40336.78 |
|  | Assault | 888333.57 | 14.6 | 42301.60 | 307548.88 | 38.4 | 38443.61 | 1195882.44 | 17.3 | 41237.33 |
| **Total** |  | **6099371.93** | **100** | **37419.46** | **801028.91** | **100** | **33376.20** | **6900400.84** | **100** | **36900.54** |

**Table 11.** Sensitivity Analysis of Estimated Lost Productivity Costs by sex, age group and cause at 6%, Cabo Verde, 2018.

| Variables |  | Male |  |  | Female |  |  | Total |  |  |
| --- | --- | --- | --- | --- | --- | --- | --- | --- | --- | --- |
|  |  | Present value | % | Mean | Present value | % | Mean | Present value | % | Mean |
| **Age group** | 15 to 19 | 20961.88 | 0.8 | 10480.94 | 20961.88 | 5.8 | 10480.94 | 41923.76 | 1.3 | 10480.94 |
|  | 20 to 24 | 301186.91 | 10.8 | 12549.45 | 37648.36 | 10.4 | 12549.45 | 338835.28 | 10.8 | 12549.45 |
|  | 25 to 29 | 281546.49 | 10.1 | 14818.24 | 74091.18 | 20.5 | 14818.24 | 355637.67 | 11.3 | 14818.24 |
|  | 30 to 34 | 309350.23 | 11.1 | 17186.12 | 0.00 | 0.0 | 0.00 | 309350.23 | 9.8 | 17186.12 |
|  | 35 to 39 | 603279.11 | 21.7 | 19460.62 | 38921.23 | 10.7 | 19460.62 | 642200.35 | 20.4 | 19460.62 |
|  | 40 to 44 | 426153.19 | 15.3 | 21307.66 | 21307.66 | 5.9 | 21307.66 | 447460.85 | 14.2 | 21307.66 |
|  | 45 to 49 | 199601.18 | 7.2 | 22177.91 | 66533.73 | 18.4 | 22177.91 | 266134.91 | 8.5 | 22177.91 |
|  | 50 to 54 | 254391.82 | 9.2 | 21199.32 | 21199.32 | 5.9 | 21199.32 | 275591.14 | 8.8 | 21199.32 |
|  | 55 to 59 | 306390.28 | 11.0 | 17021.68 | 51065.05 | 14.1 | 17021.68 | 357455.32 | 11.4 | 17021.68 |
|  | 60 to 64 | 75929.50 | 2.7 | 7592.95 | 30371.80 | 8.4 | 7592.95 | 106301.30 | 3.4 | 7592.95 |
| **Total** |  | **2778790.59** | **100** | **17047.80** | **362100.21** | **100** | **15087.51** | **3140890.80** | **100.0** | **16796.21** |
| **Causes of death** | Injuries to the head | 596240.72 | 21.5 | 17536.49 | 44075.25 | 12.2 | 14691.75 | 640315.97 | 20.4 | 17305.84 |
|  | Injuries to the neck | 15185.90 | 0.5 | 7592.95 | 0.00 | 0.0 | 0.00 | 15185.90 | 0.5 | 7592.95 |
|  | Injuries to the abdomen, lower back, lumbar spine and pelvis | 19460.62 | 0.7 | 19460.62 | 0.00 | 0.0 | 0.00 | 19460.62 | 0.6 | 19460.62 |
|  | Injuries to the hip and thigh | 0.00 | 0.0 | 0.00 | 0.00 | 0.0 | 0.00 | 0.00 | 0.0 | 0.00 |
|  | Injuries involving multiple body regions | 440134.11 | 15.8 | 16928.24 | 60740.53 | 16.8 | 15185.13 | 500874.64 | 15.9 | 16695.82 |
|  | Injuries to unspecified part of trunk, limb, or body region | 17186.12 | 0.6 | 17186.12 | 0.00 | 0.0 | 0.00 | 17186.12 | 0.5 | 17186.12 |
|  | Burns and corrosions | 38329.34 | 1.4 | 19164.67 | 35095.57 | 9.7 | 11698.52 | 73424.91 | 2.3 | 14684.98 |
|  | Poisoning by drugs, medicaments and biological substances | 17021.68 | 0.6 | 17021.68 | 0.00 | 0.0 | 0.00 | 17021.68 | 0.5 | 17021.68 |
|  | Toxic effects of substances of chiefly non-medicinal origin | 21199.32 | 0.8 | 21199.32 | 0.00 | 0.0 | 0.00 | 21199.32 | 0.7 | 21199.32 |
|  | Other and unspecified effects of external causes | 68486.66 | 2.5 | 17121.66 | 0.00 | 0.0 | 0.00 | 68486.66 | 2.2 | 17121.66 |
|  | Certain early complications of trauma not classified elsewhere | 89685.98 | 3.2 | 17937.20 | 0.00 | 0.0 | 0.00 | 89685.98 | 2.9 | 17937.20 |
|  | Accidental drowning and submersion | 287214.06 | 10.3 | 15956.34 | 0.00 | 0.0 | 0.00 | 287214.06 | 9.1 | 15956.34 |
|  | Other accidental threats to breathing | 82298.46 | 3.0 | 20574.62 | 0.00 | 0.0 | 0.00 | 82298.46 | 2.6 | 20574.62 |
|  | Intentional self-harm | 723468.81 | 26.0 | 16824.86 | 103403.77 | 28.6 | 17233.96 | 826872.58 | 26.3 | 16874.95 |
|  | Assault | 362878.81 | 13.1 | 17279.94 | 118785.09 | 32.8 | 14848.14 | 481663.90 | 15.3 | 16609.10 |
| **Total** |  | **2778790.59** | **100** | **17047.80** | **362100.21** | **100** | **15087.51** | **3140890.80** | **100** | **16796.21** |

## Discussion

This study demonstrated the socioeconomic burden due to injuries and external causes in 2018. The results of the study showed that mortality from injuries and consequences of external causes affected males disproportionately. Results indicated that 84.4% of deaths registered were among males. Similar studies have shown significantly high mortality rates in males [25–27] and these inequalities between the sexes may be associated with risky behavior, the influence of society on men’s behavior (masculinity) and in certain cases risky behavior is related to socioeconomic factors [28,29]. In this study, the mean age of mortality due to trauma was 39 years and the most affected age groups were between 20 and 44 years. The findings of this study were similar to those reported in similar studies where the active population accounted for the largest proportion of fatal victims due to injury [26,28].

The main causes of mortality by type of injury were traumatic head injuries followed by traumatic injuries involving multiple body regions. Moreover, when considering only injuries with chapter XX codes, it was observed that self-harm, assault, and drowning were the predominate causes of death. In comparison Nobre et al. and Pillay-van Wyk et al. [30,31] reported aggressions and traffic accidents as the main causes of death and while Malta et al. and Xing et al.[32,33] reported traffic accidents and suicides as the principal external causes of deah and finally, Corassa et al. [34] indicated to suicides and aggressions as the most prevalent causes. The prevalence of suicide has been associated with mental disorders and sociodemographic characteristics and aggression with socioeconomic inequalities, alcohol and drug-related harm, respectively [26,32]. In addition, traffic accidents were among the leading causes of death in Sub-Saharan Africa and while globally, traffic accidents, falls and interpersonal violence were identified as the leading causes of death due to injury [25,35].

In this analysis traffic accidents were not among the main causes of death in 2018. This was supported by data from other trauma surveillance systems that showed a low number of deaths due to traffic accidents [36]. Comprehensive road safety laws and their proper enforcement could contribute to the reduction of mortality from road accidents [25,35]. Cabo Verde approved a highway code in 2007 which has brought substantial gains in road safety [37].

In this study, assault was among the leading causes of death in the sample population and the principal cause among females. Cabo Verde has recognized gender-based violence as a social and public health problem and subsequently approved laws on the issue [38] and recognized that multiple socioeconomic and cultural factors allow the existence of this phenomenon [39]. Furthermore, gender-based violence accounted for 20% of crimes reported [36] thus, supporting the results of the current study. Insufficiency in the implementation and regulation of laws could explain the prevalence of gender based violence [34].

In the current analysis, drownings accounted for approximately 10% of mortality in the sample population, and 95.8% of these deaths were males. In Seychelles, drowning was the leading cause of death due to external causes [40]. The author associated this with the fishing industry, one of the country’s main economic and leisure activities and the need to strengthen safety measures in these industries. Despite being a small island country, similar to Cabo Verde, Abio et al. [40] suggested the promotion of swimming programs in the population as a measure to reduce the risk of drowning in the population. Drownings occurred mainly in adult males, however approximately 20% of deaths were in children in this analysis and drowning has been highlighted as one of the leading causes of death in children [34,41].

The estimated YPLL corresponded to a total of 7,222 and, a mean of 34.2 YPLL per death, which was higher than the mean obtained in Tanzania [9] and in line with the findings from Brazil, results showed that males were responsible for the highest proportion of calculated YPLL, while the age ranges from 20 to 39 were responsible for approximately 60% of YPLL [34]. The differences in the mean and prevalent age groups could explain the difference in life expectancy used to obtain YPLL and other socioeconomic factors. Similar to the current study, males contributed to a higher proportion of YPLL due to injuries, and overall injuries affect the potentially productive population [10,34,42]. Among the major contributors to YPLL were deaths due to self-harm, assault and drowning, this result corroborates findings by Delgado, (2013)[43].

Deaths between 15 and 64 years of age resulted in 4768 YPPLL, on average 25 YPPLL per death. Intentional self-harm and aggression were the main responsible for approximately 50% of YPLL and YPPLL and suicides were responsible for 30% of YPPLL. Corassa et al. [34] and Rajabali et al. [44] demonstrated that a significant proportion of YPPLL was attributable to self-harm in their studies. Additionally, injuries and other consequences from external causes accounted for 10% of lost productivity in the WHO African region, and self-harm and interpersonal violence account for 87.2% of productivity losses were due to intentional injuries [45]. In economic terms, the loss of YPLL and YPPLL in individuals aged 20 years old presented the greatest socioeconomic loss because this age corresponds to the age at which the greatest investment is made by society and the beginning of the return of that investment to society [46].

In this study, the total cost of lost productivity due to premature death from external causes was 4,580,225.91 USD men contributed 87.7% of the losses and, each death resulted in losses amounting to 24,493.19 USD. In other studies, CPL due to injury was significantly higher among men than among women [10,47]. Najafi et al. [10] reported that the highest average loss per death was due to trauma, amounting to 72,571 USD/death while Davey et al. [47] presented a mean value of approximately 2500 USD per death. A study conducted in a high-income country estimated values of lost productivity almost three times higher than the estimates found in this study [44]. Variations could be attributed to differences in the age limits, study periods, the GDP per capita and economic factors related to the countries in which the studies were conducted. The present value of the CPL at 4.5% discount rate corresponded to approximately 0.2% of GDP in 2018, considering that GDP was estimated at 1,967 billion USD. This was lower than estimates of the economic burden of road traffic accidents in low- and middle-income countries [5].

In the database used for this study, 47.1% (115) of the cases, deaths were classified by type of lesion, therefore the external causes that led to the appearance of the lesion were unknown, and 52.9% (129) were classified by external causes. ICD-10 guidelines recommended the use of codes from chapter XIX and XX to classify traumatic events and if only one code is used, chapter XX should be prioritized [17]. Results of this study demonstrate the need to standardize the classification of injuries and this could improve current estimates of mortality from external causes and complement national and global disease burden estimates [7,26]. Standardization can be achieved through regular training of health professionals on current guidelines [40] and the digitization of the registration system in health structures.

Available data on economic costs of diseases, shows that two main methods are used to evaluate the economic impact, namely the friction method and the human capital method, with the human capital method being the most used methodology in the analysis of the cost of the disease [48]. It is argued that the friction method may result more accurate estimates of lost productivity losses when compared to estimates found using the human capital method [49]. For future studies, both methods maybe applied to better evaluate the economic impact of premature mortality.

## Limitations

Data analysis confirmed the incomplete classification of causes of death due to injury. This did not allow for analysis to be done exclusively by external causes or by the anatomical site of the lesion. The approach chosen made it possible to analyze all deaths recorded due to trauma, however it made it difficult to compare with other studies on the subject. This study demonstrated the need for standardization and training in the use of recommended tools for classifying injuries. Analyzing the results presented in this study, the aforementioned limitations must be taken into account.

## Conclusion

The study demonstrated the socio-economic burden on the healthcare system due to injuries and revealed that the costs of lost productivity due to premature mortality are substantial. The analysis showed that males are disproportionately affected by trauma. In addition, the largest proportion of deaths due to injuries was in young people with the potential to contribute to the socio-economic development of the country. Attention was drawn to the prevalence and significant monetary burden of self-harm, drowning and violence in the population sample. Finally, the results of this study could be used to assess the need for implementation of other public health policies concerning the problematic of injuries, and targeted strategies for injury prevention

## Data Availability

Data cannot be shared publicly because of data transfer was not contemplated in the authorization by National Commission for Data Protection. Data are available from the National Directorate of Heath for researchers who meet the criteria for access to confidential data.

## Acknowledgment

Thanks goes to Ms. Janice Xavier, to the Board of Directors of the National Public Health Institute and the General Directorate for Planning and Budgeting of the Ministry of Health, Cabo Verde for the support given in the process of carrying out this project and to the National Directorate of Health for authorizing the use of data.

## Author’s Contributions

**Conceptualization:** Ngibo Mubeta Fernandes **Date curation:** Ngibo Mubeta Fernandes **Formal analysis:** Ngibo Mubeta Fernandes

**Financing acquisition:** Maria da Luz Lima Mendonca

**Investigation:** Ngibo Mubeta Fernandes, Lara Ferrero Gomez

**Methodology:** Ngibo Mubeta Fernandes

**Project administration:** Ngibo Mubeta Fernandes

**Supervision:** Lara Ferrero Gomez

**Resources:** Maria da Luz Lima Mendonca, Lara Ferrero Gomez

**Visualization:** Ngibo Mubeta Fernandes

**Writing - original draft:** Ngibo Mubeta Fernandes, Lara Ferrero Gomez

**Writing – proofreading and editing:** Ngibo Mubeta Fernandes, Maria da Luz Lima Mendonça, Lara Ferrero Gomez

